# Validity and Reliability of the Novel Indonesian Instrument for Aphasia Diagnosis (IDEA)

**DOI:** 10.64898/2026.07.17.26358303

**Authors:** Pukovisa Prawiroharjo, Aldithya Fakhri, Aileen Gabrielle, Violine Martalia, Sarah Alya Rahmayani, Vidya Gani Wijaya

## Abstract

Aphasia diagnosis in Indonesia remains challenging due to limited culturally and linguistically appropriate instruments. Widely used tools such as the Boston Diagnostic Aphasia Examination (BDAE) and Western Aphasia Battery (WAB) are not adapted to the Indonesian context, *while Tes Afasia untuk Diagnosis, Informasi, dan Rehabilitasi* (TADIR) provides screening but lacks diagnostic accuracy. To address this gap, we developed the *Instrumen Diagnosis dan Evaluasi Afasia* (IDEA) for native Indonesian speakers and evaluated its validity, reliability, and normative cutoff values in cognitively healthy Indonesian adults. Eighty-three cognitively normal adults (screened using MoCA-Ina) with no history of neurological disease were assessed using IDEA, which evaluates six language domains. Items were adapted from existing tools and reviewed by experts. Content validity, internal consistency (Cronbach’s alpha), and construct validity (Exploratory Factor Analysis) were analyzed using SPSS v25. A total of 83 participants were included (median age = 55.81 years, 54% secondary education). IDEA demonstrated good feasibility, with an average completion time of 45-60 minutes depending on participant engagement. Content validity was established by unanimous expert consensus. Construct validity showed meritorious sampling adequacy (KMO = .872) and significant sphericity (Bartlett’s test χ^2^ (15) = 278.523, p<.001), supporting factor analysis. Internal consistency showed good reliability across six domains (Cronbach’s α = 0.896). IDEA is a valid and reliable tool for assessing aphasia in Indonesian natives. It is a culturally appropriate assessment tool which offers structured, domain-based evaluation and supports differential diagnosis of both classical and progressive aphasia syndromes.

**Highlight:**

1. Aphasia diagnosis in Indonesia remains challenging due to limited accurate tools.
2. Our instrument evaluates language domains to assess aphasia and its mimics.
3. Validation in 83 cognitively healthy participants showed good validity and reliability.

## 1. Introduction

Aphasia is impairments in the production (motor function) and/or comprehension (sensory function) of previously normal language abilities, resulting from lesions in the dominant hemisphere of the cerebral cortex, particularly in the frontal, temporal, or parietal lobes. The most common cause of aphasia is stroke, followed by traumatic brain injury, brain tumors, and neurodegenerative diseases [1]. In Indonesia, the prevalence of language disorders due to stroke is relatively high [2]. However, a comprehensive and standardized diagnostic approach remains a major challenge in clinical practice.

One of the primary obstacles in diagnosing aphasia in Indonesia is the lack of linguistically and culturally adapted standardized assessment tools. Most available diagnostic instruments for aphasia originate from abroad and are developed in foreign languages (primarily English), such as the Boston Diagnostic Aphasia Examination (BDAE) and the Western Aphasia Battery (WAB). An Indonesian-language tool known is *Tes Afasia untuk Diagnosis, Informasi, dan Rehabilitasi* (TADIR). However, A thesis by Yugo (2022) evaluating TADIR as a diagnostic tool for aphasia concluded that, while TADIR is sensitive for screening, it lacks specificity in distinguishing aphasia subtypes and recommends the development of improved diagnostic tools with enhanced questionnaires and diagnostic algorithms [3]. This misalignment can lead to misclassification, underdiagnosis, or overdiagnosis of aphasia, and may hinder healthcare professionals in designing appropriate rehabilitative interventions [4,5].

In response to these issues, we have developed an innovative assessment tool known as the *Instrumen Diagnostik dan Evaluasi Afasia* (IDEA). IDEA is specifically designed within the linguistic and cultural context of Indonesia and evaluates various domains of language function, including thought content, speech fluency, auditory and visual comprehension, repetition, naming, reading, and writing skills. Additionally, IDEA features components to assess differential diagnoses that often accompany or mimic aphasia symptoms, such as buccofacial apraxia, verbal apraxia, ideomotor apraxia, dysarthria, paraphasia, aphasic jargon, prosopagnosia, finger agnosia, Gerstmann syndrome, and various forms of mutism [1,6]. This study aims to establish the validity, reliability, and cut-off scores of the IDEA test. These findings will provide a foundation for the next phase of research, which will assess the diagnostic accuracy of IDEA in patients with aphasia.

## 2. Materials and Methods

### 2.1. Study Design

This study employed a cross-sectional design to evaluate the validity and reliability of the *Instrumen Diagnosis dan Evaluasi Afasia* (IDEA), a novel Indonesian tool designed for aphasia diagnosis. The instrument was developed by a multidisciplinary team of neurologists and neurobehavior experts, with specific attention to the linguistic and cultural characteristics of the Indonesian language. The study was conducted following ethical approval from July 2025 to April 2026, during which participant recruitment and data collection were performed. Data analysis and manuscript preparation were completed subsequently between April 2026 and May 2026.

### 2.2. Participants

A total of 83 participants from the normal population were recruited through purposive sampling. Inclusion criteria were: (1) normal cognitive function, as determined by the Indonesian version of the Montreal Cognitive Assessment (MoCA-INA) with scores above 26, and (2) absence of neurological or psychiatric disorders. Participants with a history of brain injury, stroke, language disorder, or other medical conditions affecting cognitive function were excluded. Written informed consent was obtained from all participants prior to study enrollment.

### 2.3. Data Collection

Participants underwent a comprehensive assessment using the IDEA tool, administered by trained assessors. Data collection focused on evaluating participants’ performance across the six IDEA domains. Each session was conducted in a quiet room to minimize distractions, and responses were recorded for subsequent scoring and analysis.

### 2.4. Tools Development

The *Instrumen Diagnosis dan Evaluasi Afasia* (IDEA) was developed as a novel Indonesian-language instrument designed to comprehensively assess language and communication deficits, particularly aphasia for Indonesian native. A series of discussions and iterative item revisions were conducted to ensure that each test item was relevant, culturally appropriate, and capable of eliciting meaningful language responses from native Indonesian speakers. Content validity was assessed by expert panels, who evaluated the clarity, relevance, and representativeness of the items using a 4-point Likert scale. Items with a Content Validity Index (CVI) of <0.80 were modified or removed. The final version of IDEA underwent pilot testing on 10 cognitively normal adults, which helped to refine the task instructions, adjust the difficulty levels, and finalize the scoring guidelines.

### 2.5. Final Tool

The *Instrumen Diagnosis dan Evaluasi Afasia* (IDEA) is composed of six domains, each designed to comprehensively assess a specific aspect of language function: fluency, grammar, comprehension, naming, repetition, and writing. Fluency evaluates speech flow, phrase length, articulation, and spontaneous speech production. It includes tasks such as answering close-ended questions (LV1) and open-ended questions (LV2), generating words within a specific semantic category (LV3A), producing words beginning with a particular letter (LV3B), as well as sub-assessments for dysarthria (D1) and verbal apraxia (LV4) using familiar sentence repetition. Grammar focuses on morphosyntactic accuracy and sentence structure, with tasks that include constructing a complete sentence from three words (G1), forming a complex sentence from a single given word (G2), arranging words into grammatically correct sentences (G3), and arranging words into properly structured interrogative sentences (G4), with particular emphasis on detecting agrammatism often seen in Broca’s aphasia [7].

Comprehension is divided into verbal and written components; verbal comprehension tasks include following verbal instructions (FV1) and understanding spoken statements (FV2), while visual and written comprehension tasks involve performing actions based on written instructions (FT1) and understanding written statements (FT2), assessing both semantic and syntactic processing. Naming evaluates lexical retrieval through tasks requiring participants to name objects, activities, emotions, and places based on pictures, including confrontation naming and semantic cueing to identify anomia. Repetition measures phonological processing and working memory through normal word or sentence repetition (UV1) and tone repetition (UV2), which is specifically designed to evaluate prosody and detect dysprosody, commonly associated with right hemisphere or subcortical lesions. Finally, Writing assesses written language production through copying a paragraph (LT1), free writing (LT2), and word repetition tasks (LT3), analyzing spelling accuracy, syntax, and spontaneous written expression.

### 2.6. Statistical Analysis

Data were analyzed using SPSS version 25. Internal consistency reliability of IDEA was assessed using Cronbach’s alpha, with a value ≥0.70 considered acceptable. For validity testing, content validity index (CVI) was obtained based on expert ratings, and construct validity was examined using correlation analyses between IDEA domain scores and MoCA-INA scores. Descriptive statistics were applied to summarize participants’ demographic and test performance data.

### 2.7. Ethical Considerations

This study was approved by the Institutional Ethics Committee of Universitas Indonesia Hospital. All participants provided informed consent and were assured of confidentiality and the voluntary nature of their participation. The study was conducted in accordance with the Declaration of Helsinki.

## 3. Results

### 3.1. Demographic Characteristics

A total of 83 participants were included in this study. The demographic profile showed that there were 33 males and 50 females. The age of the participants ranged from 16 to 105 years, with the median age was 55.81 years. In terms of education level, 45 participants (54.0%) had passed secondary education or below (classified as “Low” in this study), while 38 participants (46.0%) had passed tertiary education (classified as “High”). Table 1 summarizes the demographic characteristics.

**Table 1.** Demographic Characteristics.

| **Characteristics** | **Value (n=83)** |
| --- | --- |
| **Age (median [min-max])** | 55.81 (16-105) |
| **Gender (n [%])** |  |
| Female | 50 (60) |
| Male | 33 (40) |
| **Education level** |  |
| Low (Secondary education or below) | 45 (54) |
| High (Tertiary education) | 38 (46) |

### 3.2. IDEA Instrument Performance

The descriptive statistics for the scores obtained from various components of the IDEA instrument are presented in Table 2. The sample size for each component was 83. The performance of the IDEA instrument components showed varying score ranges, means, and spread among participants. For ‘total LV %’, scores ranged from 26.62 to 100.00, with a mean of 57.00 (SD = 14.57). The 5th percentile for ‘total LV %’ was 34.00, indicating that 5% of the participants scored at or below this value. In the ‘total FT FL FV %’ component, scores varied from 54.67 to 100, averaging 85.01 (SD = 10.40). The 5th percentile for ‘total FT FL FV %’ was 63.41. ‘Total UV %’ scores fell between 60.00 and 100.00, with a mean of 93.25 (SD = 10.46), with a 5th percentile of 70.00. ‘Total N %’ scores was between 35.00 and 100.00, with a mean of 92.98 (SD = 14.38). Its 5th percentile was 65.75. For ‘total LT %’, scores ranged from 23.14 to 100, with a mean of 55.67 (SD = 18.86). The 5th percentile for ‘total LT %’ was 28.28. Finally, ‘total G %’ scores ranged from 39.07 to 82.36, with a mean of 68.86 (SD = 10.33). Its 5th percentile was 45.09.

**Table 2.** Descriptive Statistics of IDEA Instrument Components.

| Component | N | Min | Max | Mean | Std. Deviation | Percentiles 5 |
| --- | --- | --- | --- | --- | --- | --- |
| Total LV% | 83 | 26.62 | 100.00 | 57.00 | 14.57 | 34.00 |
| Total FT FL FV % | 83 | 54.67 | 100.00 | 85.01 | 10.40 | 63.41 |
| Total UV % | 83 | 60.00 | 100.00 | 93.25 | 10.46 | 70.00 |
| Total N% | 83 | 35.00 | 100.00 | 92.98 | 14.38 | 65.75 |
| Total LT% | 83 | 23.14 | 100.00 | 55.67 | 18.86 | 28.28 |
| Total G% | 83 | 39.07 | 82.36 | 68.86 | 10.33 | 45.09 |

The average time required for participants to complete the IDEA instrument ranged from 45 to 60 minutes. This variation in completion time was primarily influenced by individual participant engagement and focus during the assessment.

### 3.3. Reliability

The internal consistency of the IDEA instrument was evaluated using Cronbach’s Alpha for all six variables (total LV %, total FT FL FV %, total UV %, total N %, total LT %, total G %). The Cronbach’s Alpha coefficient for the entire scale (6 items) was 0.896. This value generally indicates good internal consistency, suggesting that the items within the instrument are moderately correlated [8].

The validity of the IDEA instrument was assessed through both content validity and construct validity. Content validity of the IDEA instrument was established through expert consensus. The instrument’s items and structure were reviewed by a panel of three experts: two medical researchers and one neurobehavioral researcher. Through internal discussions, these experts reached a unanimous agreement on the comprehensiveness, relevance, and representativeness of the instrument’s content in assessing aphasia in the Indonesian context. This expert judgment ensures that the IDEA instrument adequately covers all essential domains pertinent to aphasia diagnosis from a neurobehavioral perspective.

Construct validity was examined using exploratory factor analysis (EFA). Sampling adequacy for factor analysis was assessed using the Kaiser–Meyer–Olkin (KMO) measure, which showed a value 0.872. This indicates meritorious sampling adequacy. Bartlett’s test of sphericity was significant with approx. chi square of 278.523 (p < 0.001). These findings confirm that our data is suitable for exploratory factor analysis. The results are presented in Table 3.

**Table 3.**
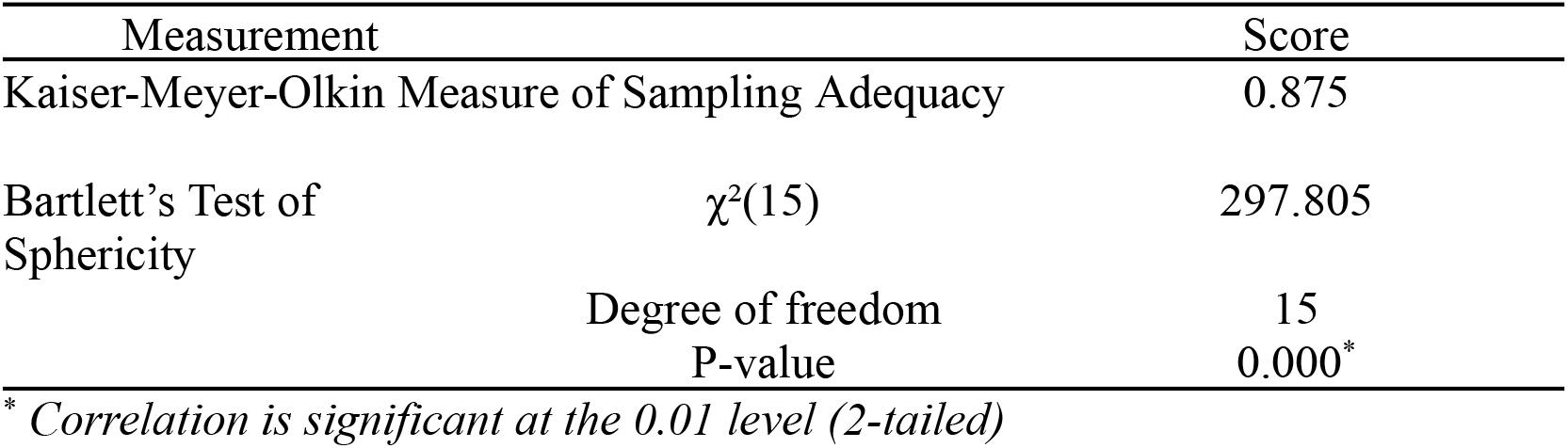
Demographic Characteristics.

## 4. Discussion

### 4.1. Comparison with Other Tools

Aphasia assessment tools play a crucial role in accurately identifying language deficits after brain injury, classifying aphasia subtypes, and determining severity to guide targeted rehabilitation. An effective tool should cover the core domains of language which includes fluency, comprehension, naming, repetition, reading, and writing, while also evaluating related cognitive and motor functions such as apraxia and prosody. Standardized scoring systems are essential to ensure reliable grading of severity and consistent subtype classification [9].

Compared with the WAB and BDAE, TADIR offers a narrower scope in assessing language and cognitive abilities. The WAB and BDAE provide a more comprehensive evaluation, encompassing apraxia, constructional tasks, detailed speech analysis, and non-verbal problem-solving. In contrast, TADIR is designed mainly to capture basic aspects of functional communication [10,11]. TADIR does not assess features such as grammatical comprehension, paraphasia types, or visual-gestural processing in detail [3].

In addition, the TADIR does not incorporate standardized scoring systems such as the Aphasia Quotient used in the WAB or the multi-dimensional indices provided by the BDAE. As a result, it offers less precision in grading severity and classifying aphasia subtypes. This limitation reduces its diagnostic depth and restricts its utility for cross-study comparisons or broader clinical applications outside the Indonesian context [3].

Subtest A of TADIR shows excellent sensitivity (97.6%), but its low specificity (21%) reduces its value as an independent diagnostic measure. While it can serve as a useful tool for initial aphasia screening, its limited ability to rule out individuals without aphasia results in a high false-positive rate. Subtest B demonstrates stronger specificity for certain subtypes, particularly transcortical sensory and global aphasia. However, its sensitivity remains inconsistent, and the positive predictive value is limited, especially for less common subtypes.^3^ These results suggest that TADIR’s structure and classification algorithm are insufficient for reliably guiding clinicians toward accurate aphasia subtype diagnoses.

IDEA builds on the gaps left by TADIR by offering a more detailed and structured approach to language assessment. Unlike TADIR, which centers on everyday communication tasks, IDEA breaks language down into six core domains: verbal fluency (LV); written language production (LT); grammar/syntax (G); written, verbal, and visual comprehension (FT FL FL); phonological repetition (UV); and lexical retrieval/naming (N). It includes features that TADIR lacks, such as screening for apraxia (subtype LV4) and dysarthria (subtype D1), as well as tasks to assess prosody, which can help detect right hemisphere or subcortical involvement (UV2). IDEA also gives greater attention to grammar and sentence construction (G1, G2, G3, G4), areas often underexplored in previous tools. Its comprehension section is divided into verbal and written tasks, allowing for more nuanced analysis. Overall, IDEA offers a fuller picture of language function and provides a stronger basis for identifying different aphasia types in clinical and research.

### 4.2. Clinical Implications

The IDEA instrument enables a structured approach to classifying secondary aphasia by systematically assessing core linguistic functions [12,13]. The diagnostic process begins with evaluating verbal fluency (LV) and written language production (LT), in line with approaches that prioritize fluency and output as initial markers of non-fluent aphasia subtypes [14]. Impairment in these domains suggests a non-fluent aphasia subtype, prompting further examination of comprehension, specifically visual (FV), written (FT), and verbal (FL), followed by phonological repetition (UV) [15,16].

Combined deficits in comprehension and repetition indicate global aphasia, while preserved repetition with impaired comprehension supports a diagnosis of mixed transcortical aphasia [7,14—18]. In contrast, if comprehension is relatively intact and repetition is impaired, the pattern corresponds to Broca’s aphasia, while preserved repetition suggests transcortical motor aphasia [13,14].

**Figure 1.**
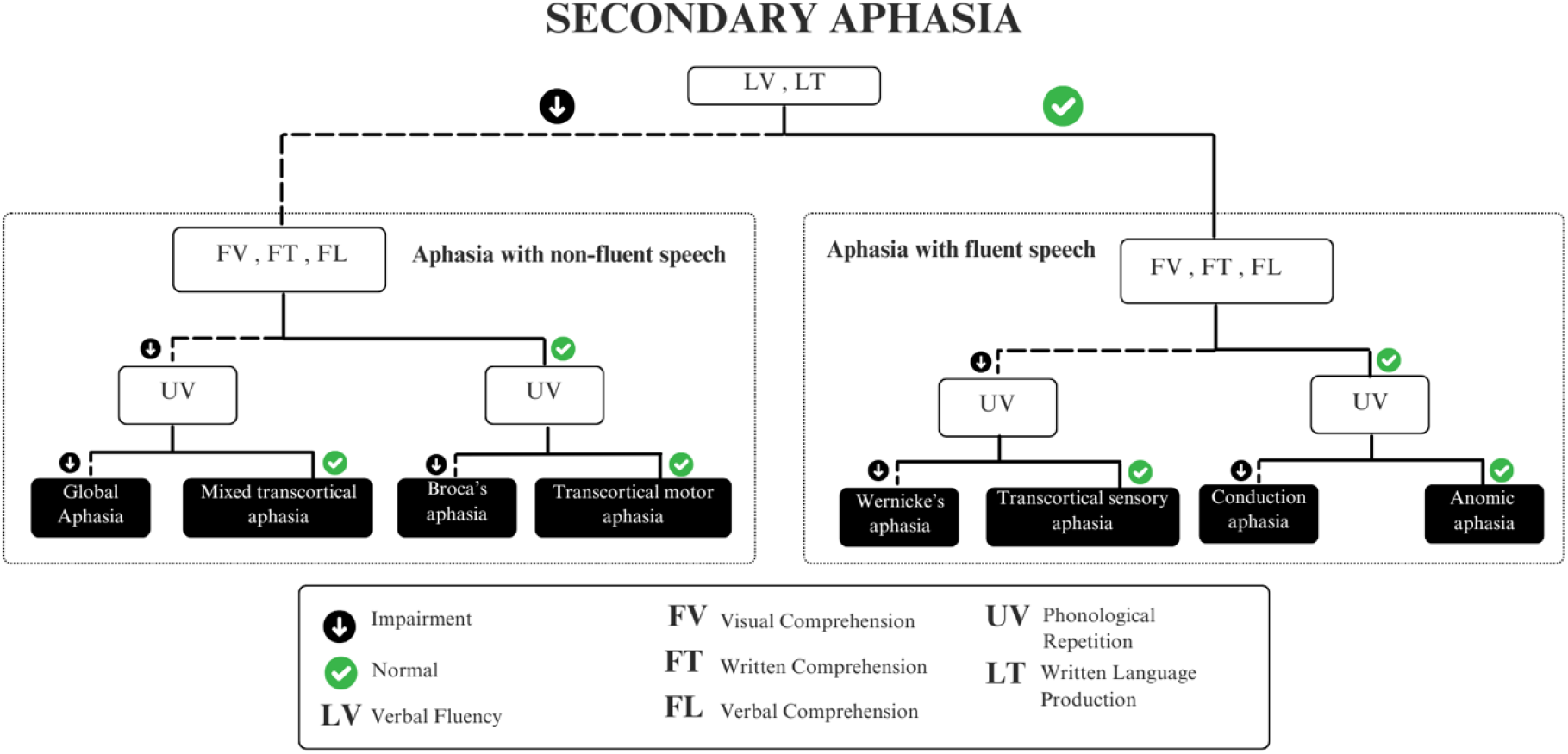
Algorithm of Secondary Aphasia using IDEA

For fluent aphasia presentations, where LV and LT are preserved, similar steps are followed. Impaired comprehension with impaired repetition indicates Wernicke’s aphasia, whereas intact repetition points to transcortical sensory aphasia. When comprehension is relatively preserved but repetition is impaired, conduction aphasia is likely. If naming deficits are predominant with otherwise preserved functions, the diagnosis is anomic aphasia [13,17,18] Through this stepwise method, IDEA offers an effective clinical pathway for identifying classical aphasia syndromes secondary to neurological insult [12,13].

The IDEA framework also supports the differentiation of primary progressive aphasia (PPA) variants by emphasizing early deficits in fluency, grammar, and comprehension [19,20]. The diagnostic sequence begins with identifying impairments in verbal fluency (LV), written language output (LT), and/or grammar and syntax (G). These markers distinguish the non-fluent variant of PPA, which can be further subclassified based on whether the primary deficit lies in syntax (agrammatic variant), motor speech production (PPA of speech), or a combination including apraxia (PPA with agrammatism and apraxia) [20,21].

**Figure 2.**
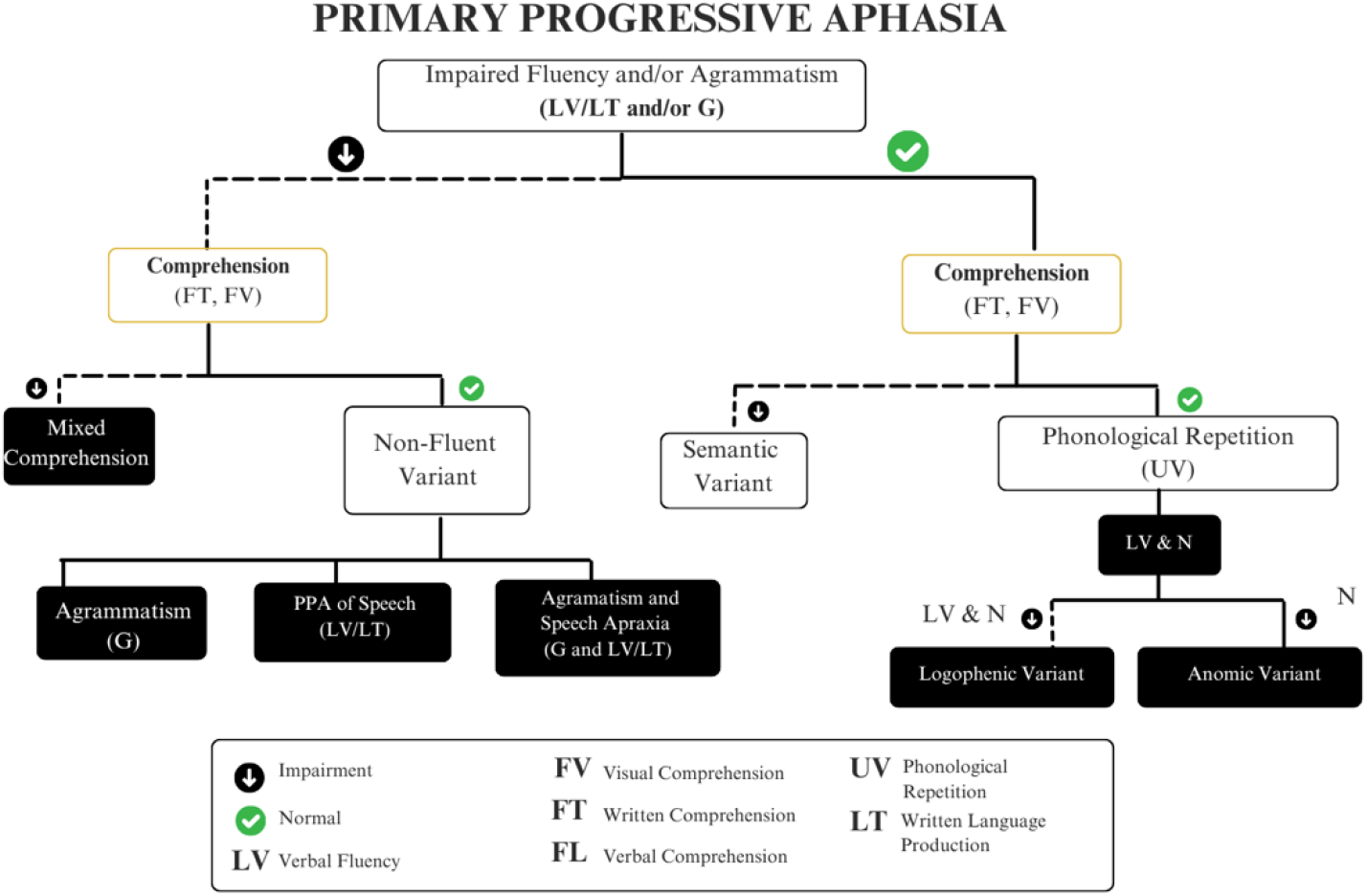
Algorithm of Primary Aphasia using IDEA

If comprehension (FT, FV) is also impaired early, the presentation is consistent with mixed comprehension variants. Conversely, if fluency and grammar are preserved but semantic comprehension deteriorates, particularly in object and word meaning, the diagnosis shifts to semantic variant PPA [22,23]. Patients with intact comprehension and repetition (UV) but impaired naming (N), with or without fluency disturbance, fall under either the logopenic variant (both LV and N impaired) or anomic variant (only N impaired) [20,21]. IDEA’s comprehensive evaluation of linguistic and cognitive-linguistic domains enables refined, syndrome-specific diagnosis in PPA, reflecting current diagnostic criteria while maintaining clinical feasibility.

### 4.3. Limitations and Future Directions

One key limitation of this study lies in the sampling distribution, particularly regarding participant age. Although efforts were made to recruit a representative sample, the age groups were not proportionately distributed. This imbalance may limit the generalizability of the instrument’s validity and reliability across different age cohorts [24,25]. Given that cognitive, emotional, or physical factors influencing responses can vary with age, the results may not fully capture the performance of the instrument across the lifespan [26,27]. In addition, age-related familiarity with certain visual stimuli used in the instrument, such as images depicting electronic devices, or images depicting emotions using emoticons, may have influenced response accuracy. Future research should aim to put these factors into consideration to ensure balanced representation across age groups and therefore strengthen the external validity of the findings.

## 5. Conclusions

The *Instrumen Diagnosis dan Evaluasi Afasia* (IDEA) is a valid and reliable tool for assessing aphasia in Indonesian-speaking populations. It offers a culturally adapted, domain-based approach with strong content and construct validity. The use of percentile cutoffs and a structured diagnostic flowchart enhances its clinical applicability in identifying both classical and progressive aphasia syndromes.

## Declarations

### Ethical Approval Statement

This study was approved by the Institutional Ethics Committee of Universitas Indonesia Hospital (protocol code: S-183/KETLIT/RSUI/VII/2025, accepted July 25^th^ 2025). All participants provided informed consent and were assured of confidentiality and the voluntary nature of their participation. The study was conducted in accordance with the Declaration of Helsinki.

### Data Availability Statement

The datasets generated and/or analyzed during this study are available from the corresponding author upon reasonable request.

### Author Contributions

Conceptualization, P.P., A.F., A.G., V.M., S.A.R.; Methodology, P.P., A.F., A.G., V.M., S.A.R., V.G.; Sampling, P.P., A.F., A.G., V.M., S.A.R., V.G.; Results analysis, A.G., A.F., V.M. S.A.R.; Writing—original draft preparation, P.P., A.F., A.G., V.M., S.A.R., V.G.; Writing—review and editing, P.P., A.F., A.G., V.M., S.A.R., V.G.; Visualization, A.G, S.A.R.; Supervision, P.P.; Project administration, P.P., A.F., A.G., V.M., S.A.R., V.G. All authors have read and agreed to the published version of the manuscript.

### Informed Consent Statement

Informed consent was obtained from all subjects involved in the study.

## Acknowledgments

The authors would like to acknowledge the experts and clinicians who provided valuable insights and recommendations during the development of the *Instrumen Diagnosis dan Evaluasi Afasia* (IDEA) tool. The authors would also thank the participating hospitals, healthcare facilities, and the patients who generously contributed their time to participate in this study.

## References

[1] H. Le, F. Lui, M.Y. Lui Aphasia, in: StatPearls, StatPearls Publishing, Treasure Island, FL, 2026.

[2] H.N. Tasari, M. Muryanti, Efektivitas metode semantik divergen terhadap kemampuan bahasa ekspresif pada penderita afasia lancar di Kecamatan Jebres [Effectiveness of divergent semantic methods on expressive language ability in patients with fluent aphasia in Jebres District], J. Ter. Wicara Bahasa. 1 (2023) 289–296. 10.59686/jtwb.v1i2.41.

[3] Y.P. Hardito, P. Prawiroharjo, R. Yetty, L.N. Diatri, K. Mohammad, Uji diagnostik tes afasia untuk diagnosis, informasi, dan rehabilitasi (TADIR) sebagai instrumen penegakan diagnosis dan tipe afasia, [Diagnostic test of aphasia for diagnosis, information, and rehabilitation (TADIR) as an instrument for determining diagnosis and aphasia type], Master’s Thesis, Faculty of Medicine Universitas Indonesia, 2022.

[4] P.M. Pedersen, K. Vinter, T.S. Olsen, Aphasia after stroke: type, severity and prognosis, Cerebrovasc. Dis. 17 (2004) 35–43. 10.1159/000073896.

[5] S.M. Sheppard, R. Sebastian, Diagnosing and managing post-stroke aphasia, Expert Rev. Neurother. 21 (2021) 221–234. 10.1080/14737175.2020.1855976.

[6] K. Morihara, S. Ota, K. Kakinuma, N. Kawakami, Y. Higashiyama, S. Kanno, et al., Buccofacial apraxia in primary progressive aphasia, Cortex. 158 (2023) 61–70. 10.1016/j.cortex.2022.10.010.

[7] E. Fedorenko, R. Ryskin, E. Gibson, Agrammatic output in non-fluent, including Broca’s, aphasia as a rational behavior, Aphasiology. 37 (2023) 1981–2000. 10.1080/02687038.2022.2143233.

[8] M. Tavakol, R. Dennick, Making sense of Cronbach’s alpha, Int. J. Med. Educ. 2 (2011) 53–55. 10.5116/ijme.4dfb.8dfd.

[9] M.W.M. Fong, R. Van Patten, R.P. Fucetola, The factor structure of the Boston Diagnostic Aphasia Examination, Third Edition, J. Int. Neuropsychol. Soc. 25 (2019) 772–776. 10.1017/S1355617719000237.

[10] F.I. Fitri, A.S. Rambe Gustianingsih, D. Widayati, Clinical and linguistic profiles and challenges in diagnosis of primary progressive aphasia in Medan, Indonesia: a hospital-based study, Open Neurol. J. 18 (2024) e1874205X305965. 10.2174/011874205X305965240607112722.

[11] Z. Yan, D. Wei, S. Xu, J. Zhang, C. Yang, X. He, et al., Determining levels of linguistic deficit by applying cluster analysis to the aphasia quotient of Western Aphasia Battery in post-stroke aphasia, Sci. Rep. 12 (2022) 15108. 10.1038/s41598-022-17997-0.

[12] A.D. Halai, B. De Dios Perez, J.D. Stefaniak, M.A. Lambon Ralph, Efficient and effective assessment of deficits and their neural bases in stroke aphasia, Cortex. 155 (2022) 333–346. 10.1016/j.cortex.2022.07.014.

[13] K. Berg, J. Isaksen, S.J. Wallace, M. Cruice, N. Simmons-Mackie, L. Worrall, Establishing consensus on a definition of aphasia: an e-Delphi study of international aphasia researchers, Aphasiology. 36 (2022) 385–400. 10.1080/02687038.2020.1852003.

[14] K. Monnelly, J. Marshall, L. Dipper, M. Cruice, A systematic review of intensive comprehensive aphasia programmes—who takes part, what is measured, what are the outcomes?, Disabil. Rehabil. 46 (2024) 4335–4349. 10.1080/09638288.2023.2274877.

[15] S.D. Teti, L.L. Murray, J.B. Orange, K.S. Kankam, A.C. Roberts, Telepractice assessments for individuals with aphasia: a systematic review, Telemed. e-Health. 31 (2025) 37–49. 10.1089/tmj.2024.0268.

[16] L. Hammond, T. Christensen, J. Fridriksson, D.B. den Ouden, Assessing functional communication in persons with aphasia: a scoping review of formal and informal measures, Int. J. Lang. Commun. Disord. 60 (2025) e70051. 10.1111/1460-6984.70051.

[17] D.S. Kasselimis, P.G. Simos, C. Peppas, I. Evdokimidis, C. Potagas, The unbridged gap between clinical diagnosis and contemporary research on aphasia: a short discussion on the validity and clinical utility of taxonomic categories, Brain Lang. 164 (2017) 63–67. 10.1016/j.bandl.2016.10.005.

[18] L. Springer, S. Mantey, The Comprehensive Aphasia Test: a review. Commentary on Howard, Swinburn, and Porter, “Putting the CAT out: What the Comprehensive Aphasia Test has to offer”, Aphasiology. 24 (2010) 75–78.

[19] M.L. Gorno-Tempini, A.E. Hillis, S. Weintraub, A. Kertesz, M. Mendez, S.F. Cappa, et al., Classification of primary progressive aphasia and its variants, Neurology. 76 (2011) 1006–1014. 10.1212/WNL.0b013e31821103e6.

[20] D.C. Tippett, Classification of primary progressive aphasia: challenges and complexities, F1000Res. 9 (2020) 64. 10.12688/f1000research.21184.1.

[21] P. Hoffman, S.A. Sajjadi, K. Patterson, P.J. Nestor, Data-driven classification of patients with primary progressive aphasia, Brain Lang. 174 (2017) 86–93. 10.1016/j.bandl.2017.08.001.

[22] L. Fernandez-Romero, J. Carrick, R. Landin-Romero, D. Foxe, M. Yus-Fuertes, A. Marcos-Dolado, et al., Cognitive profiles in primary progressive aphasia variants: a cross-cultural Australian and Spanish investigation, J. Neurol. Sci. 472 (2025) 123446. 10.1016/j.jns.2025.123446.

[23] M.M. Mesulam, C. Wieneke, C. Thompson, E. Rogalski, S. Weintraub, Quantitative classification of primary progressive aphasia at early and mild impairment stages, Brain. 135 (2012) 1537–1553. 10.1093/brain/aws080

[24] M. Piccininni, J.L. Rohmann, M. Wechsung, G. Logroscino, T. Kurth, Should cognitive screening tests be corrected for age and education? Insights from a causal perspective, Am. J. Epidemiol. 192 (2023) 93–101. 10.1093/aje/kwac159.

[25] O. Hatahet, M.L. Seghier, The validity of studying healthy aging with cognitive tests measuring different constructs, Sci. Rep. 14 (2024) 23880. 10.1038/s41598-024-74488-0.

[26] T.S. Marks, G.M. Giles, D.F. Edwards, Functional cognitive performance augments cognitive screening data in older adults, Front. Aging Neurosci. 17 (2025) 1535146. 10.3389/fnagi.2025.1535146.

[27] M.A. Heiskanen, J. Nevalainen, K. Pahkala, M. Juonala, N. Hutri, M. Kähönen, et al., Change in cognitive performance during seven-year follow-up in midlife is associated with sex, age, and education—The Cardiovascular Risk in Young Finns Study, J. Neurol. 271 (2024) 5165–5176. 10.1007/s00415-024-12466-2.

